# THE INDONESIA HEALTH WORKFORCE QUANTITY AND DISTRIBUTION

**DOI:** 10.1101/2024.03.31.24305126

**Authors:** Farizal Rizky Muharram, Hanif Ardiansyah Sulistya, Julian Benedict Swannjo, Fikri Febrian Firmansyah, Muhammad Masrur Rizal, Alifina Izza, Muhammad Atoillah Isfandiari, Ninuk Dwi Ariningtyas, Achmad Chusnu Romdhoni

## Abstract

**Background:** Indonesia, the world’s largest archipelago, faces unique challenges in distributing its health workforce across its diverse geographic barriers, leading to disparities in health worker number and distribution. By dissecting the distribution patterns and identifying areas of critical need, the research seeks to inform policy interventions that can more effectively bridge the gap on health worker quantity and inequity.

**Methods:** We conducted a descriptive analysis of healthcare workforce data across all 514 districts in Indonesia. The study focused on five categories of health workers: General Practitioners (GPs), medical specialists, dentists, nurses, and midwives. We calculated the health worker ratio to determine the availability of healthcare workers relative to the population. To evaluate the distribution of these workers, we employed the Gini Index as a measure of distribution equality. Additionally, we conducted a comparative metric approach to assess both the quantity and the equity of healthcare worker distribution across the districts.

**Results:** In Indonesia, the current health worker ratio stands at 3.84 per 1000 population, falling short of the WHO’s threshold of 4.45 for achieving 80% Universal Health Coverage. This shortfall translates to a need for an additional 166,000 health workers. Our analysis reveals a varied distribution of health worker categories: while midwives show a relatively equitable distribution, specialists and dentists exhibit significant inequality, especially at the district level. The Gini Index, used to measure this inequality, indicates greater disparities at the district level compared to the provincial level. There has been notable progress in the distribution of medical specialists across provinces, with the between-provinces Gini Index for specialists decreasing from 0.57 in 1993 to 0.44 in 2022. However, the inter-district Gini Index remains high at 0.53 in 2022, signifying a concentration of specialists in major cities and provincial capitals.

**Conclusion:** This study shows that human resources for health in Indonesia suffer not only in quantity but also in distribution. Our finding underscores the importance of considering inter-province and inter-district disparities to tailor policies to tackle unique problems each region faces.

**Evidence Before Study:**

- Prior research has established that the quantity and distribution of health workers are critical factors in improving life expectancy and are fundamental components of the health system.
- Following Indonesia’s constitutional changes in 2001, which included the autonomy and decentralization of healthcare services, assessing the number of health workers at the district level has gained significant importance for determining national healthcare needs.
- There has been a notable gap in studies analyzing the quantity and distribution of health workers in Indonesia’s district level. Previous research often missed the nuances of district-level challenges, focusing instead on broader, national-level assessments.

**What This Study Adds:**

- **First National Study on Health Workforce:** This is the first study of health workforce quantity and distribution at Indonesia’s National level. so this paper serves as a basic reference for future research
- **Quantitative Analysis of Distribution Equity:** Utilizing the Gini Index, the study quantifies the level of inequality in the distribution of healthcare workers, offering a clear metric to guide policymakers in assessing and addressing regional disparities.
- **Identification of Regional Variations:** The study highlights significant regional variations, with some provinces showing an inequitable distribution of health workers, demonstrating a critical need for increased healthcare personnel and better distribution strategies.
- **Dual-dimensional assessment:** The study introduces a quadrant comparative approach that simultaneously evaluates the quantity of healthcare workers and the equity of their distribution across Indonesian provinces. This dual-dimensional analysis is a significant methodological advancement, providing a more holistic understanding of healthcare workforce allocation.

**How This Study Might Affect Research, Practice, and/or Policy:** This study has the potential to become the basis of policy-making related to the distribution of health workers and provide constructive feedback and strategical insights that could be utilized to decrease the gap between health workers and their maldistribution.

## INTRODUCTION

The landscape of global health is continually evolving, marked by the dynamic interplay between healthcare needs and the availability of healthcare professionals. At the heart of this landscape is the pivotal role of health workers, whose quantity and distribution are essential for effective healthcare delivery^1^. The World Health Organization (WHO) since 2005 and aligned with Sustainable Development Goals (SDGs) indicators underscore this significance, particularly after adjusting the need of universal health coverage ^2,3^. Human resources for health, serving as the backbone of health systems, face the persistent challenge of achieving equitable distribution. With its unique geographical and demographic profile, Indonesia exemplifies this struggle, presenting a complex case for the distribution and availability of health workers.

Indonesia, the world’s largest archipelago and fourth most populous nation, with its approximately 17,000 islands, embodies these challenges in distributing its health workforce^4^. The country’s vast and varied geography, encompassing remote areas, borders, and small islands, complicates the equitable distribution of healthcare services^5^. Additionally, the decentralized migration of health workers towards urban centers exacerbates the issue, leading to a disparity in healthcare availability between urban and rural or remote regions^6^ .

The number of health workers is crucial in determining the effectiveness of healthcare delivery^7^. However, the issue extends beyond mere numbers; the distribution of these workers plays a critical role in ensuring that all regions, irrespective of their geographic or demographic characteristics, have access to adequate healthcare services. This challenge is further compounded by the issue of healthcare workforce production in Indonesia, which is not accompanied by equitable distribution ^8–10^. The imbalance in the distribution of health workers, particularly in a geographically diverse country like Indonesia, presents a multifaceted challenge that needs comprehensive analysis and targeted interventions ^11^.

The aim of this study is to provide an in-depth analysis of the current state of health workers in Indonesia, with a particular focus on both their quantity and distribution at the district level. Previous reports and research have often overlooked broader regional or national-level assessments^12^. This lack of detailed data has been a significant gap in understanding the true extent of the challenge faced by the Indonesian health system. By delving into the district-level dynamics of health worker distribution and quantity, this study seeks to bridge this knowledge gap and offer insights that could inform policy and practice, ultimately contributing to reducing inequities in healthcare access across Indonesia.

## METHODS

We gather information from various sources, particularly formal government agencies and ministries that collect data on health resources, both published and unpublished. All the analysis needed will be done in the R using stats and the dineq package.

### Definition of Administrative Unit

Indonesia’s administrative area is divided into 34 provinces and 514 districts. Districts are categorized as cities (urban areas) or regencies(rural areas), with 416 districts and 98 cities reported in 2022. Health worker data is also aggregated into five major islands: Sumatra, Java, Kalimantan, Bali-Nusa, Sulawesi, and Maluku-Papua. This approach considers that health service coverage can extend beyond district and province administrative boundaries.

### Definition and Source of Health Worker Data

Health workers are general practitioners, specialists, dentists, nurses, and midwives. Health workers must have gone through higher education and professional training^13,14^. This study consolidates all 24 categories of specialists into a single group. General practitioners (GPs) practice both in primary and referral care, and between private and public. while specialists primarily operate within hospitals and private practices. Midwives are predominantly employed in primary care.

Number of health workers at the provincial-level data is derived from the *“Annual Indonesian Health Profile Book”* from 1990 to 2021. District-level data were gathered from the *“Minister of Health’s Health Resources Database (SISDMK)”.* We extracted SISDMK data on 31 December 2022. Data on doctors, dentists, and specialists are supplemented with Data on Members of the Indonesian Doctors Association and Doctors’ Registration Data by the Indonesian Medical Council. The Health worker ratio will represent the number of health workers per 1,000 population within a geographic unit. Population estimate data, used as the health worker ratio’s denominator, were retrieved from the Ministry of Internal Affairs’ GIS data, with the most recent estimates dated 30 June 2022.

### Benchmark of Health Worker Density

The existing density of health workers will be compared with goals set by Indonesia’s Directorate of Health Workforce Planning’s Target Document, utilizing Data Envelopment Analysis (DEA) and Stochastic Frontier Analysis (SFA) for target ratio calculations (see Table 1). We will also compared to the World Health Organization (WHO) proposed threshold of health-worker ratio (including doctors, nurses, and midwives) of 4.45 per 1,000 people to meet 80% coverage of the Sustainable Development Goals (SDGs)^15^

**Table 1.**
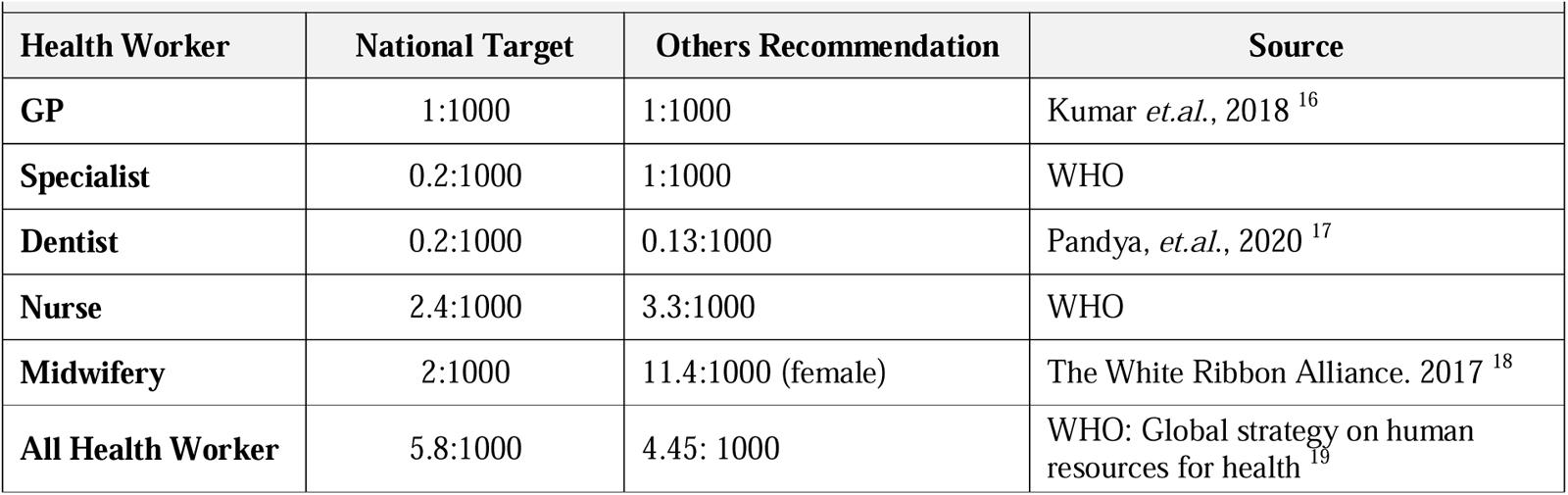
Recommendation of Health Worker density.

### Distribution Analysis

To analyze distribution, we will directly compare ratios between administrative, urban, and rural areas and use the Gini Index, as previously recommended in WHO Measuring Health Worker inequalities^20^ ^21–23^. The Gini coefficient is a statistical metric obtained from the Lorenz curve, which measures the level of inequality in a distribution. It is expressed as a value between 0 (indicating perfect equality) and 1 (indicating perfect inequality). We performed inter-provincial and within-province Gini Index, which has been prevalently used in prior research ^24–27^. To further measure inequities between district, the inter-district Gini index will be used in this research^28,29^

### Quadrant Analysis Framework

We made a quadrant analysis to evaluate the two facets of quantity and distribution, as depicted in Figure 1. This method involves plotting provinces on a two-dimensional plane defined by two axes: the Gini Index and the Health Worker (HW) Ratio. The quadrant framework categorizes provinces into four distinct groups based on their position relative to the median values of the Gini Index and HW Ratio. The resulting quadrants are labeled as follows:

1. **Scarce and Inequitable Province**: High Gini Index and Low HW Ratio, indicating a province with a shortage of health workers and a highly uneven distribution.
2. **Inequitable Province**: High Gini Index and High HW Ratio, suggesting a province with a sufficient number of health workers but an unequal distribution among the population.
3. **Scarce Province**: Low Gini Index and Low HW Ratio, pointing to a province with an overall scarcity of health workers but a relatively even distribution among the population.
4. **Ideal Province**: Low Gini Index and High HW Ratio, representing a province with both a high number of health workers and an equitable distribution.

**Figure 1.**
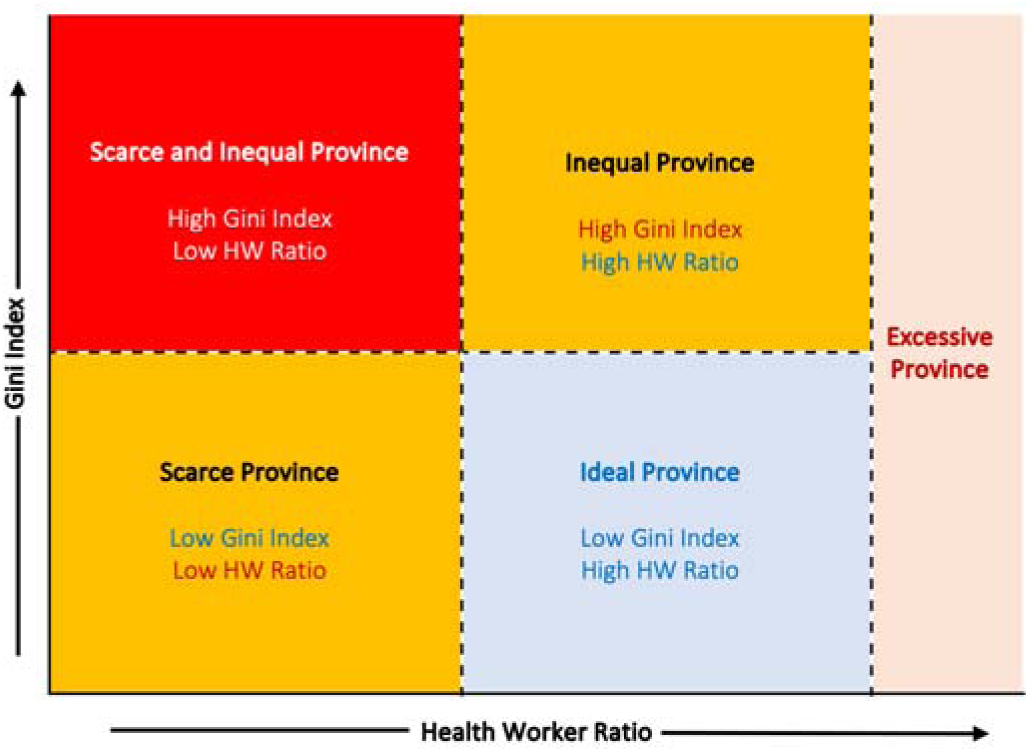
A Quadrant Analysis of Gini Index vs. Health Worker Ratio

This framework is intended to identify and visualize the disparities in health worker distribution across provinces. In line with the nature of Indonesia, decentralization pushes the provincial government to achieve equal growth within the province. This approach will inform policymakers at the provincial and national levels by highlighting areas of need and guiding targeted interventions.

## RESULTS

### Quantity of Health Workers

Indonesia’s health workforce comprises 1,046,640, averaging 3.84 per 1,000 people nationally but varying widely across districts (0.31 to 19.08). Nurses are the largest group (531,214; 1.95 per 1,000), followed by midwives (300,756; 1.10 per 1,000), general practitioners (145,602; 0.38 per 1,000), specialists (41,893; 0.15 per 1,000), and dentists (27,177; 0.1 per 1,000). The nurse-to-doctor ratio is about 2.17 nationally.

In examining the distribution of health workers between urban and rural areas in Indonesia, clea disparities are evident across different medical professions. General Practitioners (GPs) in rural areas have a median ratio of 0.27 and an average of 0.31, contrasting sharply with urban areas, where the median ratio is substantially higher at 1.10 and the average is 1.34. The disparity is even more pronounced for dentists; in rural regions, they have a median ratio of 0.05 and an average o 0.06, whereas in urban areas, these figures significantly increase to a median of 0.20 and an average of 0.27. Specialists present a similar pattern of distribution. In rural areas, the median ratio for specialists is 0.06 with an average of 0.08, which is markedly lower than in urban areas, where the median ratio rises to 0.35 and the average to 0.42.

Rural areas have median ratios of 0.87 for midwives and 1.26 for nurses, which are substantially lower compared to urban areas, where the median ratios are 0.86 for midwives and a significantly higher 3.18 for nurses. Overall, this data underscores a consistent trend of higher health worker For most healthcare worker categories, the distribution is right-skewed, meaning that most rural and urban areas have lower ratios of healthcare workers, with a few areas having much higher ratios. The kurtosis values suggest that these distributions are leptokurtic, with a higher peak and heavier tails than a normal distribution, particularly for GPs and dentists in rural areas and for urban nurses.

**Table 2.**
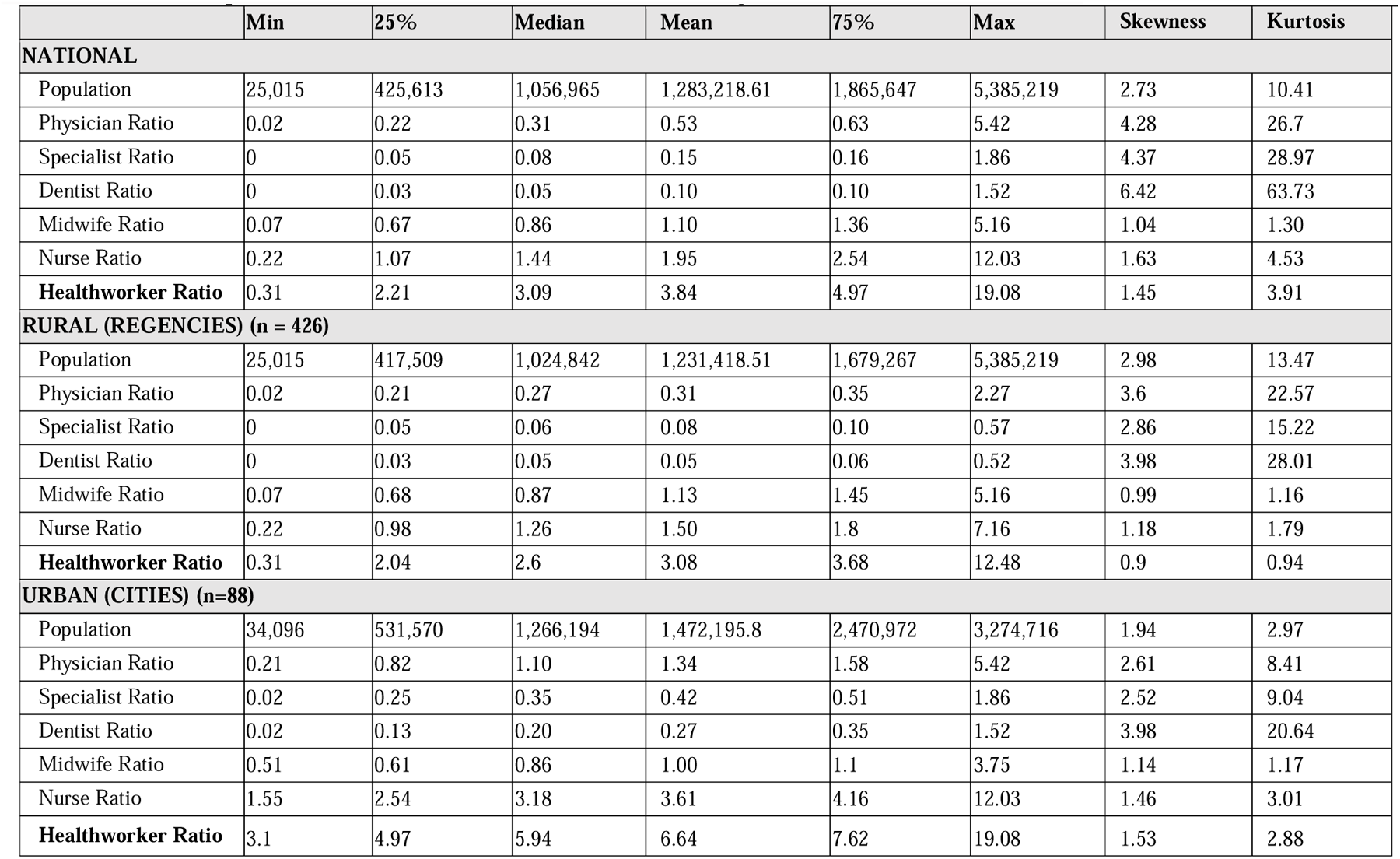
Descriptive Statistics of Health workers density in Indonesia.

### Island and Provincial Levels Distribution

Table 3 shows the distribution of health worker ratios across Indonesian islands and provinces. The data reveals several disparities, with Java having the lowest health worker ratio (3.06 per 1,000 population) and Sulawesi the highest (5.04 per 1,000 population). Each island presents a distinct health worker composition; notably, Bali Nusa Tenggara and Java have the highest GP ratios at 0.45 and 0.38, respectively. Java leads in specialist and dentist ratios (0.17 and 0.1) but has the lowest ratios for nurses and midwives (1.69 and 0.72), suggesting an imbalance in healthcare worker distribution.

Provincially, Aceh has the highest health worker ratio (7.68), followed by DKI Jakarta (6.23). The lowest ratios are in West Java (2.44), Banten (2.69), and Central Java (2.83). Bali’s GP ratio (0.98) is close to the WHO-recommended physician ratio, and DKI Jakarta has the highest specialist ratio (0.63). These figures indicate uneven healthcare resource allocation, with certain areas having a high concentration of healthcare professionals and others facing severe shortages.

**Figure 2.**
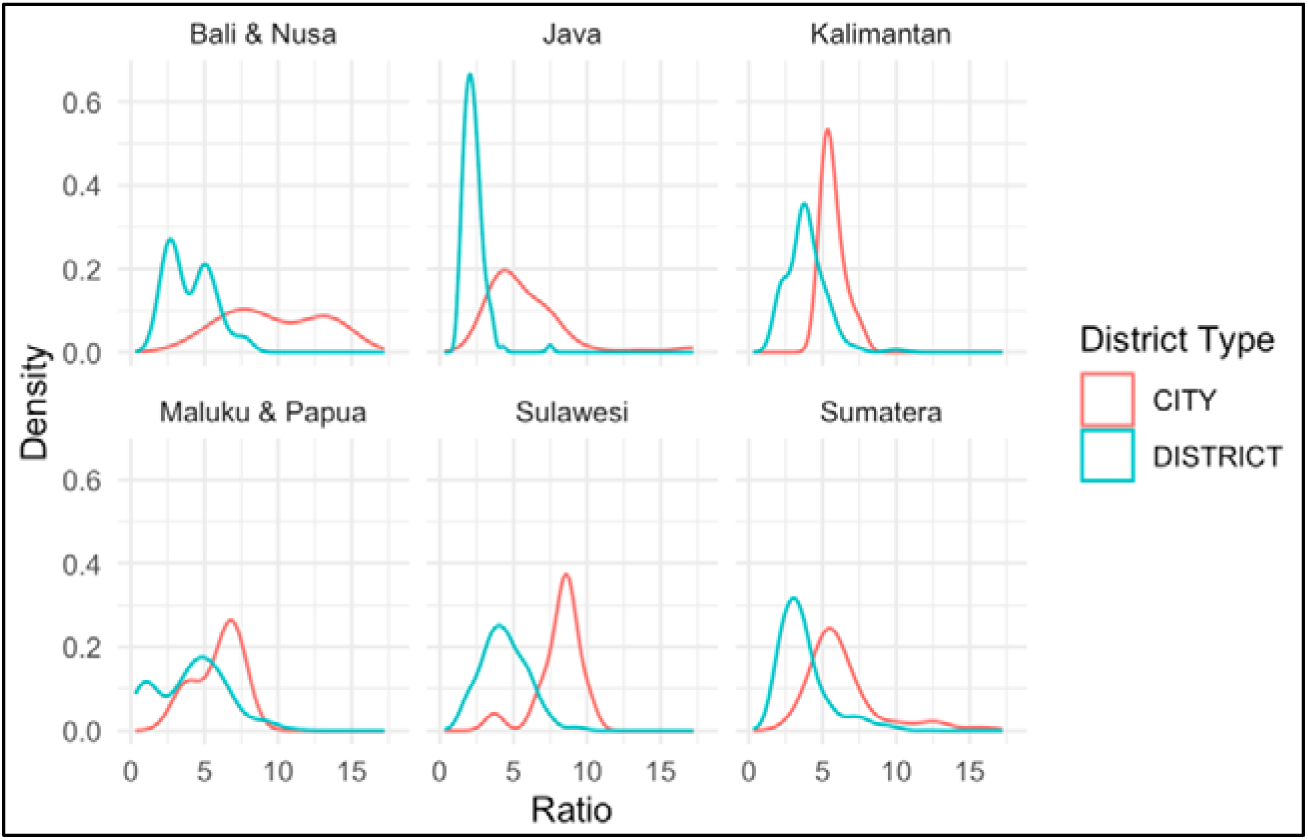
Regional Health Worker Distribution by City and District: A Density Plot Comparison. This set of density plots compares the distribution of health worker ratios across different Indonesian regions, distinguishing between city and district types. Peaks represent the most common ratios within each region, with red lines indicating city densities and blue lines for districts. The plots reveal variations in distribution patterns, with some regions showing distinct differences between city and district densities, potentially indicating disparities in health worker allocation.

The density plot at figure 2 presents a comparative analysis of health worker (HW) ratios across different islands and regions in Indonesia, categorized by city and district types. In Bali and Nusa, the HW ratio distribution for cities and districts demonstrates a bimodal pattern, indicating two distinct groupings of HW ratios within each category. Java’s distribution exhibits a stark peak for cities at a lower HW ratio, suggesting a uniformity in lower ratios among cities, in contrast to districts that show a broader, more varied distribution.

Kalimantan’s data suggests a single, similar peak for both cities and districts, with cities having a marginally higher peak, implying a slight concentration of HW ratios within city areas. Meanwhile, Maluku & Papua reveal a significant contrast between cities and districts; cities have a flatter, more varied HW ratio distribution, whereas districts show a distinct, narrow peak at higher ratios. Sulawesi’s density plot indicates a single peak for both city and district types, with the city peak being more pronounced, pointing towards a more consistent HW ratio among Sulawesi cities.

Sumatera’s analysis reveals a higher peak for districts compared to cities, suggesting a trend towards higher HW ratios within district areas. Overall, the graph indicates significant regional variability in HW ratios, with certain trends such as the bimodal distribution in Bali & Nusa and the contrasting distributions in Maluku & Papua standing out.

**Table 3.**
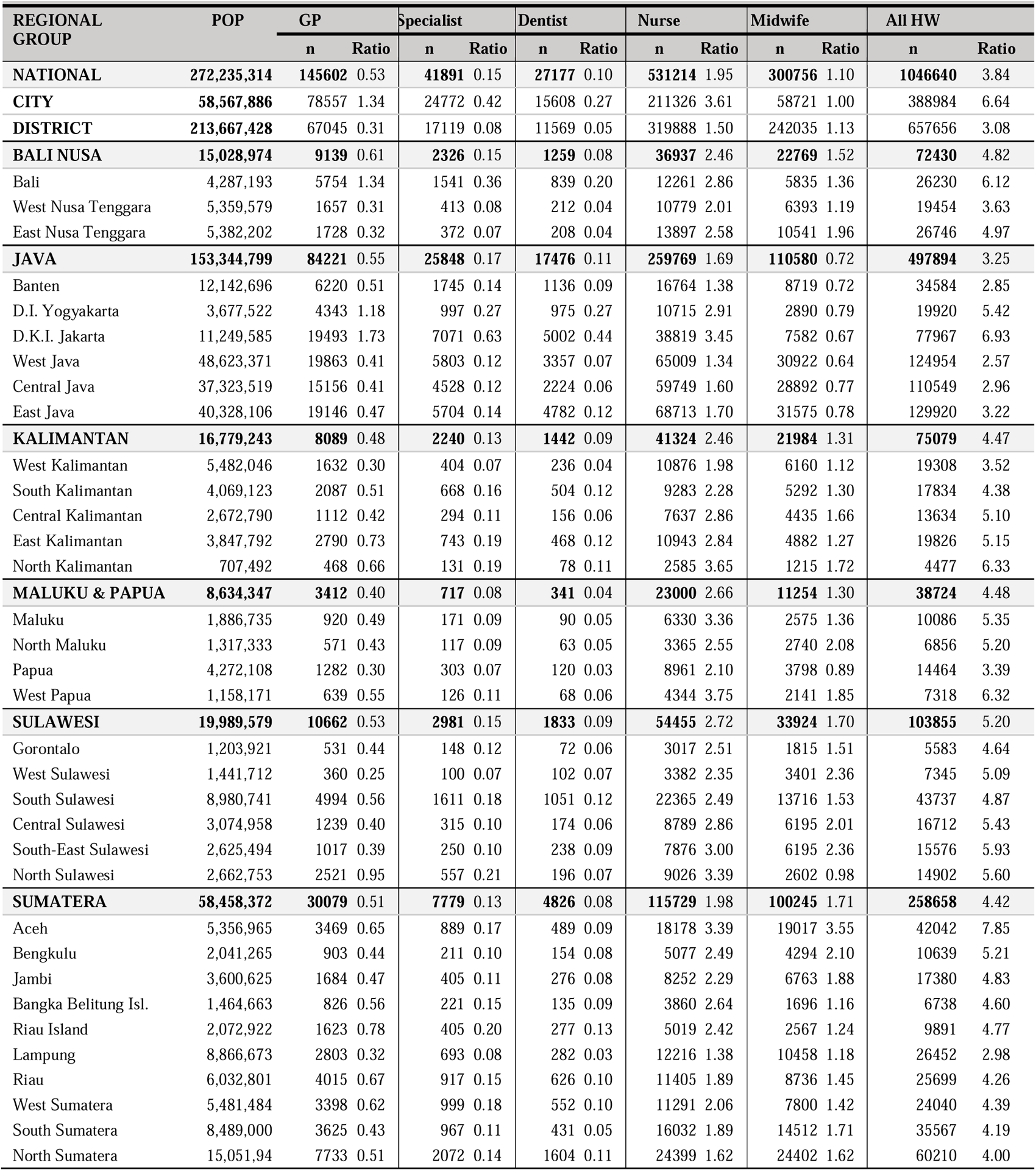
Healthcare Workforce Distribution by Region: Population-to-Health Worker Ratios in Indonesia. The ratios are calculated per 1,000 population.

| REGIONAL GROUP | POP | GP |  | Specialist |  | Dentist |  | Nurse |  | Midwife |  | All HW |  |
| --- | --- | --- | --- | --- | --- | --- | --- | --- | --- | --- | --- | --- | --- |
|  |  | n | Ratio | n | Ratio | n | Ratio | n | Ratio | n | Ratio | n | Ratio |
| **NATIONAL** | **272,235,314** | **145602** | 0.53 | **41891** | 0.15 | **27177** | 0.10 | **531214** | 1.95 | **300756** | 1.10 | **1046640** | 3.84 |
| **CITY** | **58,567,886** | 78557 | 1.34 | 24772 | 0.42 | 15608 | 0.27 | 211326 | 3.61 | 58721 | 1.00 | 388984 | 6.64 |
| **DISTRICT** | **213,667,428** | 67045 | 0.31 | 17119 | 0.08 | 11569 | 0.05 | 319888 | 1.50 | 242035 | 1.13 | 657656 | 3.08 |
| **BALI NUSA** | **15,028,974** | **9139** | 0.61 | **2326** | 0.15 | **1259** | 0.08 | **36937** | 2.46 | **22769** | 1.52 | **72430** | 4.82 |
| Bali | 4,287,193 | 5754 | 1.34 | 1541 | 0.36 | 839 | 0.20 | 12261 | 2.86 | 5835 | 1.36 | 26230 | 6.12 |
| West Nusa Tenggara | 5,359,579 | 1657 | 0.31 | 413 | 0.08 | 212 | 0.04 | 10779 | 2.01 | 6393 | 1.19 | 19454 | 3.63 |
| East Nusa Tenggara | 5,382,202 | 1728 | 0.32 | 372 | 0.07 | 208 | 0.04 | 13897 | 2.58 | 10541 | 1.96 | 26746 | 4.97 |
| **JAVA** | **153,344,799** | **84221** | 0.55 | **25848** | 0.17 | **17476** | 0.11 | **259769** | 1.69 | **110580** | 0.72 | **497894** | 3.25 |
| Banten | 12,142,696 | 6220 | 0.51 | 1745 | 0.14 | 1136 | 0.09 | 16764 | 1.38 | 8719 | 0.72 | 34584 | 2.85 |
| D.I. Yogyakarta | 3,677,522 | 4343 | 1.18 | 997 | 0.27 | 975 | 0.27 | 10715 | 2.91 | 2890 | 0.79 | 19920 | 5.42 |
| D.K.I. Jakarta | 11,249,585 | 19493 | 1.73 | 7071 | 0.63 | 5002 | 0.44 | 38819 | 3.45 | 7582 | 0.67 | 77967 | 6.93 |
| West Java | 48,623,371 | 19863 | 0.41 | 5803 | 0.12 | 3357 | 0.07 | 65009 | 1.34 | 30922 | 0.64 | 124954 | 2.57 |
| Central Java | 37,323,519 | 15156 | 0.41 | 4528 | 0.12 | 2224 | 0.06 | 59749 | 1.60 | 28892 | 0.77 | 110549 | 2.96 |
| East Java | 40,328,106 | 19146 | 0.47 | 5704 | 0.14 | 4782 | 0.12 | 68713 | 1.70 | 31575 | 0.78 | 129920 | 3.22 |
| **KALIMANTAN** | **16,779,243** | **8089** | 0.48 | **2240** | 0.13 | **1442** | 0.09 | **41324** | 2.46 | **21984** | 1.31 | **75079** | 4.47 |
| West Kalimantan | 5,482,046 | 1632 | 0.30 | 404 | 0.07 | 236 | 0.04 | 10876 | 1.98 | 6160 | 1.12 | 19308 | 3.52 |
| South Kalimantan | 4,069,123 | 2087 | 0.51 | 668 | 0.16 | 504 | 0.12 | 9283 | 2.28 | 5292 | 1.30 | 17834 | 4.38 |
| Central Kalimantan | 2,672,790 | 1112 | 0.42 | 294 | 0.11 | 156 | 0.06 | 7637 | 2.86 | 4435 | 1.66 | 13634 | 5.10 |
| East Kalimantan | 3,847,792 | 2790 | 0.73 | 743 | 0.19 | 468 | 0.12 | 10943 | 2.84 | 4882 | 1.27 | 19826 | 5.15 |
| North Kalimantan | 707,492 | 468 | 0.66 | 131 | 0.19 | 78 | 0.11 | 2585 | 3.65 | 1215 | 1.72 | 4477 | 6.33 |
| **MALUKU & PAPUA** | **8,634,347** | **3412** | 0.40 | **717** | 0.08 | **341** | 0.04 | **23000** | 2.66 | **11254** | 1.30 | **38724** | 4.48 |
| Maluku | 1,886,735 | 920 | 0.49 | 171 | 0.09 | 90 | 0.05 | 6330 | 3.36 | 2575 | 1.36 | 10086 | 5.35 |
| North Maluku | 1,317,333 | 571 | 0.43 | 117 | 0.09 | 63 | 0.05 | 3365 | 2.55 | 2740 | 2.08 | 6856 | 5.20 |
| Papua | 4,272,108 | 1282 | 0.30 | 303 | 0.07 | 120 | 0.03 | 8961 | 2.10 | 3798 | 0.89 | 14464 | 3.39 |
| West Papua | 1,158,171 | 639 | 0.55 | 126 | 0.11 | 68 | 0.06 | 4344 | 3.75 | 2141 | 1.85 | 7318 | 6.32 |
| **SULAWESI** | **19,989,579** | **10662** | 0.53 | **2981** | 0.15 | **1833** | 0.09 | **54455** | 2.72 | **33924** | 1.70 | **103855** | 5.20 |
| Gorontalo | 1,203,921 | 531 | 0.44 | 148 | 0.12 | 72 | 0.06 | 3017 | 2.51 | 1815 | 1.51 | 5583 | 4.64 |
| West Sulawesi | 1,441,712 | 360 | 0.25 | 100 | 0.07 | 102 | 0.07 | 3382 | 2.35 | 3401 | 2.36 | 7345 | 5.09 |
| South Sulawesi | 8,980,741 | 4994 | 0.56 | 1611 | 0.18 | 1051 | 0.12 | 22365 | 2.49 | 13716 | 1.53 | 43737 | 4.87 |
| Central Sulawesi | 3,074,958 | 1239 | 0.40 | 315 | 0.10 | 174 | 0.06 | 8789 | 2.86 | 6195 | 2.01 | 16712 | 5.43 |
| South-East Sulawesi | 2,625,494 | 1017 | 0.39 | 250 | 0.10 | 238 | 0.09 | 7876 | 3.00 | 6195 | 2.36 | 15576 | 5.93 |
| North Sulawesi | 2,662,753 | 2521 | 0.95 | 557 | 0.21 | 196 | 0.07 | 9026 | 3.39 | 2602 | 0.98 | 14902 | 5.60 |
| **SUMATERA** | **58,458,372** | **30079** | 0.51 | **7779** | 0.13 | **4826** | 0.08 | **115729** | 1.98 | **100245** | 1.71 | **258658** | 4.42 |
| Aceh | 5,356,965 | 3469 | 0.65 | 889 | 0.17 | 489 | 0.09 | 18178 | 3.39 | 19017 | 3.55 | 42042 | 7.85 |
| Bengkulu | 2,041,265 | 903 | 0.44 | 211 | 0.10 | 154 | 0.08 | 5077 | 2.49 | 4294 | 2.10 | 10639 | 5.21 |
| Jambi | 3,600,625 | 1684 | 0.47 | 405 | 0.11 | 276 | 0.08 | 8252 | 2.29 | 6763 | 1.88 | 17380 | 4.83 |
| Bangka Belitung Isl. | 1,464,663 | 826 | 0.56 | 221 | 0.15 | 135 | 0.09 | 3860 | 2.64 | 1696 | 1.16 | 6738 | 4.60 |
| Riau Island | 2,072,922 | 1623 | 0.78 | 405 | 0.20 | 277 | 0.13 | 5019 | 2.42 | 2567 | 1.24 | 9891 | 4.77 |
| Lampung | 8,866,673 | 2803 | 0.32 | 693 | 0.08 | 282 | 0.03 | 12216 | 1.38 | 10458 | 1.18 | 26452 | 2.98 |
| Riau | 6,032,801 | 4015 | 0.67 | 917 | 0.15 | 626 | 0.10 | 11405 | 1.89 | 8736 | 1.45 | 25699 | 4.26 |
| West Sumatera | 5,481,484 | 3398 | 0.62 | 999 | 0.18 | 552 | 0.10 | 11291 | 2.06 | 7800 | 1.42 | 24040 | 4.39 |
| South Sumatera | 8,489,000 | 3625 | 0.43 | 967 | 0.11 | 431 | 0.05 | 16032 | 1.89 | 14512 | 1.71 | 35567 | 4.19 |
| North Sumatera | 15,051,94 | 7733 | 0.51 | 2072 | 0.14 | 1604 | 0.11 | 24399 | 1.62 | 24402 | 1.62 | 60210 | 4.00 |

### District-Level Distribution

Figure 3 shows a distinct change in the mapping of health workers between the provincial-level aggregate ratio(1a) and district-level aggregate ratio(1b), especially in Java Island. Health workers are concentrated in cities (small areas with a high ratio of health workers), and lack of health workers in more rural areas in Java. Despite having the largest number of healthcare workers, by comparing the number to their population, the ratio in most districts in Java remained below Indonesia and WHO-recommended density (Figure 4).

**Figure 3.**
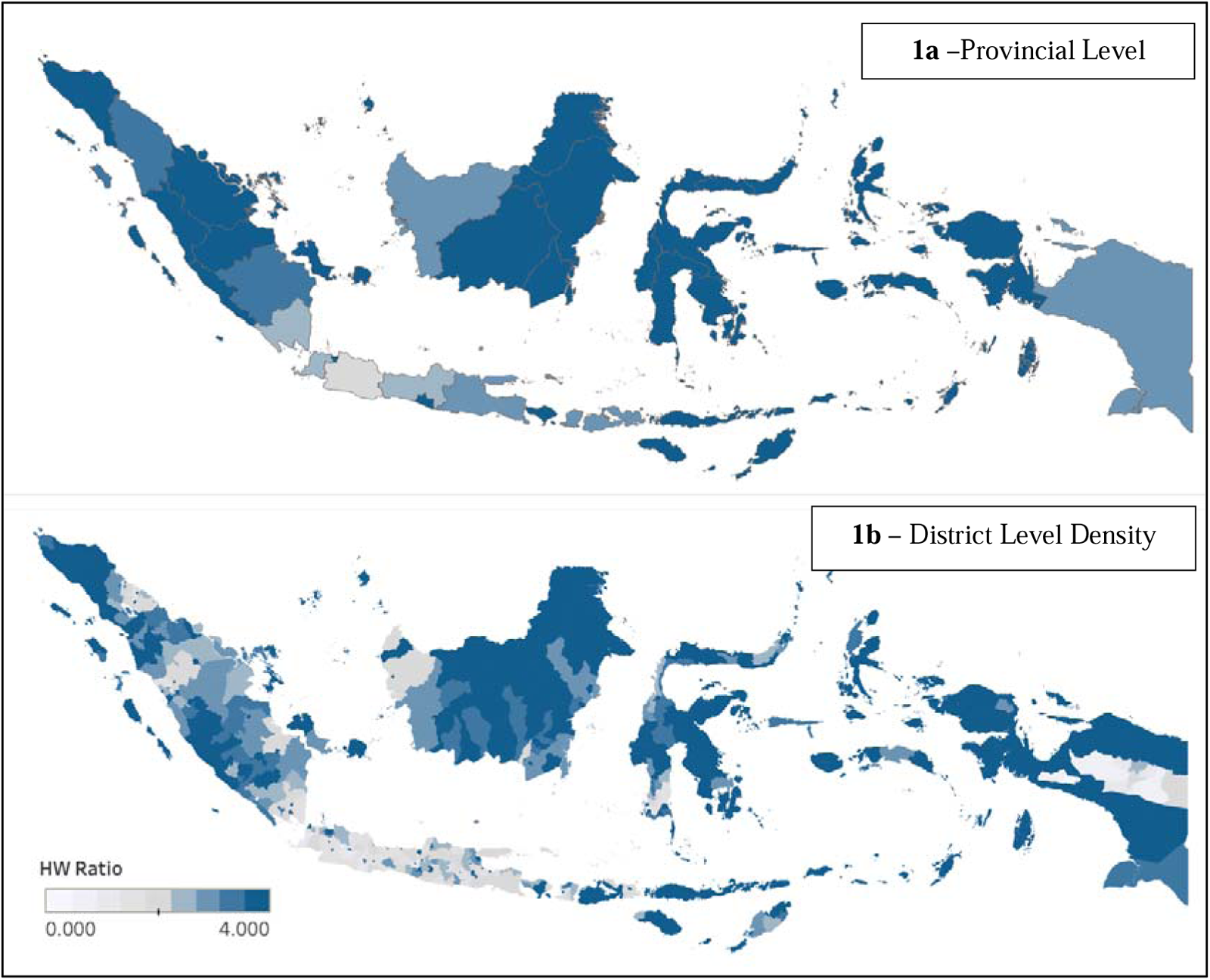
Geographic Distribution of Health Worker Ratios in Indonesia: A Comparative View at Provincial and District Levels. This map pair illustrates the health worker (HW) ratio across Indonesia’s provinces (1a) and districts (1b). Darker shades of blue indicate higher HW ratios, with the scale ranging from 0 to 4. The provincial-level map shows broader trends, while the district-level map provides a more granified perspective, revealing significant variations within provinces.

**Figure 4.**
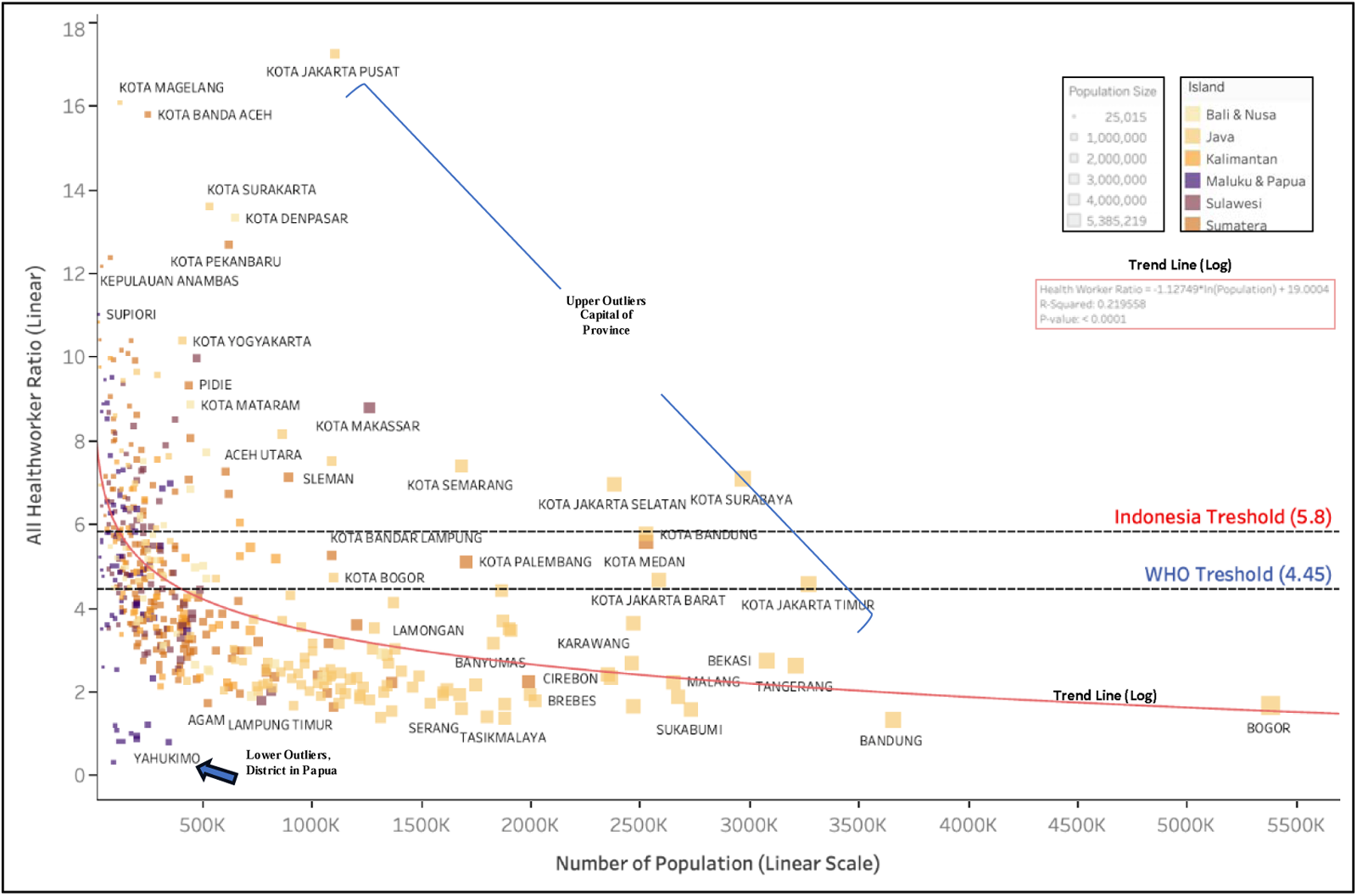
Population vs. Health Worker Ratio Across Indonesian Regions: Scaling and WHO Standards Compliance. The scatter plot displays the relationship between the logarithm of the population size and the health worker ratio for various Indonesian regions. Each point represents a region, with the WHO-recommended ratio as a reference line. The plot highlights disparities in health worker distribution relative to population, with some regions significantly above or below the WHO benchmark.

The Figure 4 scatter plot displays a wide variance in health worker-to-population ratios. The graph indicates that most regions have health worker ratios below the density target except for a few districts that are also the province’s capital. Several regions, particularly in Papua, such as Yahukimo, are identified as lower outliers with a deficiency in health worker density.

Districts with a larger population tend to have fewer health workers. A logarithmic trend line fitted to the data demonstrates a decremental trend, suggesting that regions with larger populations tend to have a lower health-worker ratio. The Logarithmic trend line equation quantifies the relationship between the population size and the health worker ratio, Health Worker Ratio = - 1.2749*ln(Population) + 19.004. The coefficient of determination, R-squared (0.215958). Moreover, the distribution of health worker ratios exhibits right-skewness. This skewness indicates that while most regions are understaffed, a few are exceptionally well-staffed.

### Inequity in Distribution

The Gini Index has demonstrated variable trends over the past three decades, as depicted in Figure 4 and Table 4. For General Practitioners (GPs), the provincial Gini Index increased from 0.35 in 1992 to a peak of 0.49 in 2003 before declining to 0.39 in 2012 and rising again to 0.42 in 2022. This fluctuation suggests changes in the equality of GP distribution over time. Dentists and nurses also experienced an increase in provincial inequality, with their Gini Indexes rising to 0.49 and 0.40, respectively, in 2022. Midwives, however, showed an improvement in equality from 1992 to 2022, with the Gini Index decreasing from 0.38 to 0.35.

**Table 4.** Provincial and District-Level Inequality in Health Worker Distribution: Gini Index from 1992 to 2022. This table presents the Gini Index scores for different health worker categories, measuring the inequality in their distribution across provinces from 1992 to 2022 and at the district level in 2022. Data Unavailability is marked as ’…’

| Health worker | Between Province Gini Index |  |  |  |  |  |  | Between District Gini Index 2022 |
| --- | --- | --- | --- | --- | --- | --- | --- | --- |
|  | 1992 | 1997 | 2003 | 2008 | 2012 | 2017 | 2022 |  |
| GP | 0.35 | 0.45 | 0.49 | 0.42 | 0.39 | 0.39 | 0.42 | 0.46 |
| Specialist | 0.57 | 0.57 | ... | ... | 0.42 | 0.45 | 0.44 | 0.53 |
| Dentist | 0.38 | 0.43 | 0.41 | 0.42 | 0.40 | 0.43 | 0.49 | 0.52 |
| Nurse | 0.34 | 0.36 | 0.37 | ... | 0.36 | 0.38 | 0.40 | 0.33 |
| Midwife | 0.38 | 0.38 | 0.39 | 0.39 | 0.41 | 0.36 | 0.35 | 0.31 |

Disparities at the district level, recorded for 2022, reveal a more pronounced inequality for health workers than at the provincial level. GPs saw their district-level Gini Index escalate to 0.46, dentists to 0.52, and specialists to the highest at 0.53. In contrast, Nurses and midwives exhibited a reduced inequality at the district level, with a Gini Index of 0.33 and 0.31, respectively, signifying a more between-province inequality rather than relatively equal distribution across districts.

**Figure 5.**
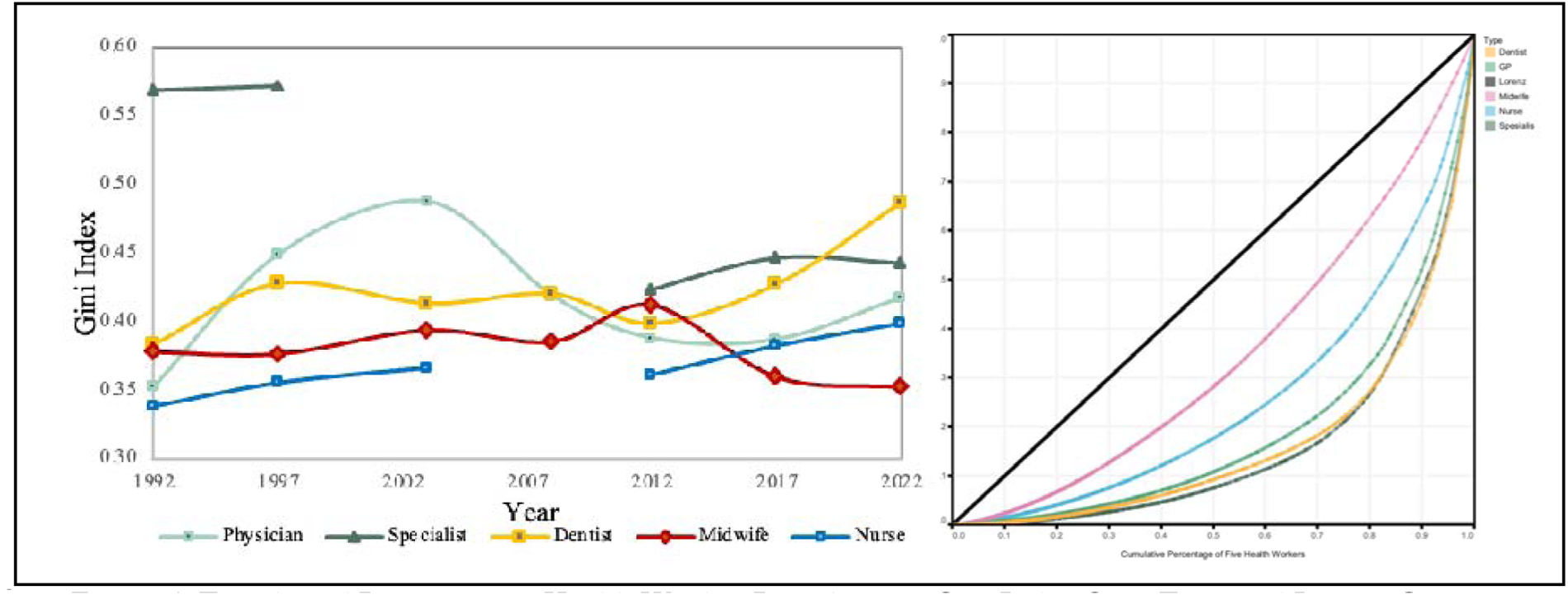
Trends and Disparities in Health Worker Distribution: Gini Index Over Time and Lorenz Curve Analysis. The graph comprises two panels: the left shows the Gini Index for different health worker over 30 years, indicating inequality levels within each sector. The right panel’s Lorenz Curves visualize the actual distribution equity for these professions Each color represents a distinct healthcare role, ensuring consistency across both visual analyses.

Supported by the Lorenz curves, it suggests that the bottom percentage of districts have significantly fewer health workers than would be expected in an equal distribution, especially for GPs, specialists, and dentists. The middle of the distribution for these categories also shows a considerable gap from the line of equality, indicating that the Gini index’s sensitivity to the middle distribution reflects the reality of the distribution of health workers. The top district(s) seem to have a disproportionately high number of health workers, especially specialists and dentists, as indicated by the curves’ steep rise as they approach 100%.

### Scarcity and Inqeuity

Figure 6 offers a visual evaluation of health worker distribution across Indonesian regions. The y-axis is the Gini Index to assess equity in distribution against the x-axis as health worker ratios for various categories, including general practitioners, specialists, dentists, midwives, and nurses.

The health worker scatterplot illustrates a widespread distribution across the provinces, with a noticeable lean towards lower ratios, which suggests a general scarcity of health workers in many areas. North Sumatra, in particular, registers a high Gini Index coupled with a low health-worker ratio, underscoring an urgent need for redistribution and an increase in the health workforce.

The scatterplots for physicians, specialists, and dentists display a similar pattern, with most provinces failing to meet minimum health worker targets. With exceptions such as Jakarta, Yogyakarta, and Bali that exceed these targets. With its five districts and a substantial physician population, Jakarta is uniquely situated in the bottom-right quadrant, highlighting an oversupply of physicians compared to other regions clustered to the left. **S**pecialists are predominantly located in the upper-left quadrant, indicating a skewed distribution that favors urban centers or provincial capitals and suggests a concentration of specialists in fewer, more developed areas. With Nurses exhibit the most equitable distribution, with only Papua showing significant disparities. However, the range between the highest and lowest ratios remains considerable, from 1.2 to 3.6 pe 1,000. The Midwives ratio plot identifies Aceh as the only province exceeding the equity threshold. Conversely, Jakarta, typically lower-right in other health worker categories, deviates in the case o Midwive, showing a surprisingly low ratio and positioning it towards the left quadrant.

**Figure 6.**
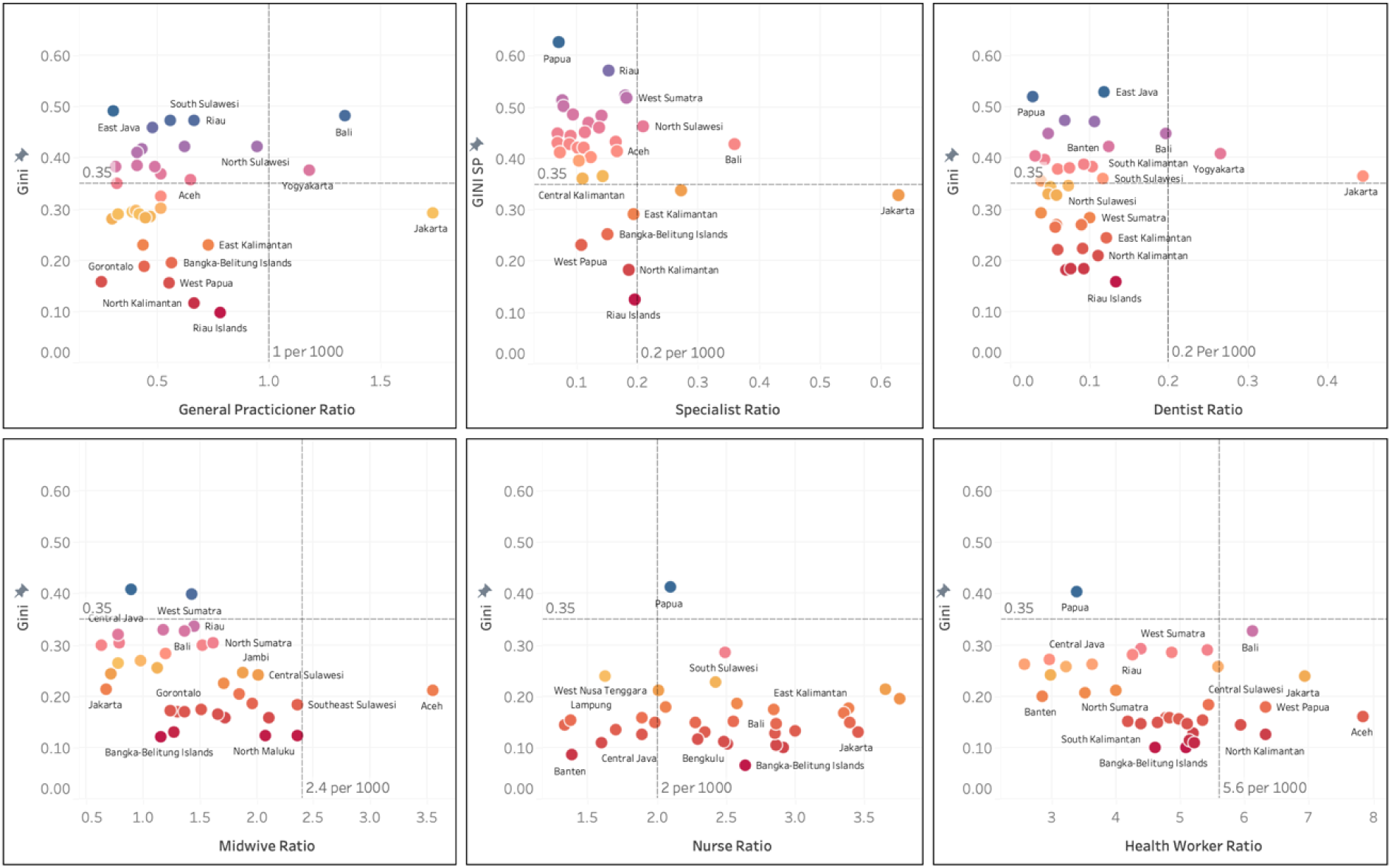
Comparative Analysis of Health Worker Distribution Across Regions: Gini Index vs. Various Health Worker Ratios. (Each scatter plot represents a different category of health workers, with the Gini index on the vertical axis and the respective health worker ratio on the horizontal axis. Do represents a province, with blue shades indicating regions with a higher Gini of health. The dashed line for Gini referenced 0.35 Gini Ratio, the target for equality. Dashed line for Ratio referenced to national Target for each health worker)

## DISCUSSION

### Quantity of Health Workers

Indonesia’s archipelagic geography, with its varied terrain and areas isolated by mountains and oceans, adds to the distribution challenges. Among its 80,000 villages, over 20,000 face geographical difficulties that obstruct access to healthcare. As of 2022, Indonesia’s health-worker ratio is 3.84. The threshold set by WHO to achieve 80% Universal Health Coverage is 4.45— requiring an additional 166,000 health workers to surpass the 4.45 threshold.^30^ Indonesia’s lack o active GPs and Specialists requires attention (0.64 physicians per 1,000 population). Low compared to other LMIC Nations such as India (0.73 per 1000), China (2.4 doctors per 1,000), and other ASEAN Countries (Singapore (2.29), the Philippines (0.6), and Malaysia (1.54).) ^31^. Indonesia mus add up to 100,000 physicians to meet the threshold.^17–20^

### Decomposition of Inequity

Contrary to the general perception of Java being saturated with health workers. By numbers, Java has the most healthcare workers, but Java has the lesser density of total health workers, mainly caused by the low number of nurses and midwives. Java is the only island with a midwives ratio lower than 1 and a nurse ratio lower than 2 per 1000 population. However, Java is also the island with the highest ratio of specialists (0.17), while Maluku and Papua had a ratio lower than half of Java’s (0.8).

Most islands have nearly reached the WHO threshold, with only Java (3.52) and Sumatra (4.42) still below it. However, the high population on these islands significantly affects the national average of health workers. None of the islands have yet met the Indonesian Government’s threshold of 5.8 health workers per 1000 population, with Sulawesi coming closest at 5.2.

The key issue lies in the composition of health workers. Java faces a shortage of nurses and midwives despite having a relatively high number of doctors, though still below the threshold. In contrast, islands outside of Java have a relative abundance of nurses and midwives but a scarcity of doctors.^32^ This imbalance in the distribution of health workers can be addressed through aggressive policy measures and enhanced nurse education, as demonstrated by China’s experience. In China, around 2003, the ratio of nurses to physicians was quite low. However, recent data indicates a significant improvement, with the ratio rising from 1 nurse per 1000 population in 2003 to 4.1 nurses per 1000 population in 2018.^33^

There is a profound disparity between urban and rural areas. Up to 50% of the total GP was in the City, despite the city population being only 20% of the total Population. That’s also happening in India, where Out of all qualified workers, 77.4% were located in urban areas, even though the urban population is only 31% of the country’s total population. Cities exhibit higher mean ratios across all healthcare workforce categories. Like other low- and lower-middle-income nations, the city tends to have a more significant health worker density.^23,34–42^ While still skewed, the midwife and nurse ratios are less severe.^43^ This finding is consistent with other research that found an uneven distribution of the total health workforce between rural and urban areas.^44–46^

### Inequity Indicator

The overall trend indicates that while some health worker categories, like midwives, have achieved an equal distribution, others, particularly specialists and dentists, face higher levels of inequality, especially considering the district level. This discrepancy is highlighted by the Gini Index differences between the provincial and district levels, with an apparent increase in inequality at the finer scale of districts for most health worker categories.

There has been some progress in equalizing the distribution of medical specialists across provinces. The between-provinces Gini Index For Specialists remarkably dropped from 0.57 in 1993 to 0.44 in 2022. However, the Inter-district Gini Index is very high at 0.53 in 2022. This implies that specialists are still saturated in the preferable district, huge cities, and the province’s capital.^34,47,48^ Measuring only the between-province Gini Index will mask the disparities within provinces and interdistricts.

Comparative studies from western China, a less affluent region compared to the eastern and southern parts of the country, exhibit similar inequality trends. The Physician Gini Index here stands at 0.46. The World Health Organization’s last published data from 2010 shows relatively more minor interdistrict disparities in China, with a Gini Index of 0.362 for doctors and 0.471 for nurses. In India, these figures are 0.436 for doctors and 0.527 for nurses^49,50^. The situation in Indonesia contrasts with both China and India, as it exhibits a higher Gini Index for general physicians and specialists than nurses.

### Two Faces of Health Worker Problem in Indonesia: Inequity and Scarcity

The quadrant analysis provides a multidimensional view of healthcare workforce distribution across regions. It highlights the areas of need and the complexity of issues like scarcity and inequality that often overlap, making it difficult to discern the underlying causes and appropriate interventions.

The upper-right quadrant has a high ratio of health workers, but resources are not evenly spread across the province. Bali serves as an illustrative case within this analysis. For instance, urban centers like Denpasar City might be well-staffed, while more remote areas suffer from a dearth of healthcare professionals. Addressing this imbalance requires policies incentivizing healthcare workers to relocate to underserved areas or increasing the training and recruitment of health workers specifically for these regions.

The upper-left quadrant represents a more critical scenario, as seen with East Java and Papua regions. The inequality in their distribution worsens the simultaneously general scarcity of health workers. Such provinces struggle with redistribution because the overall number of health workers is too low to meet even the minimum needs. To mitigate this, a two-pronged approach is essential: boosting the healthcare workforce numbers through training and recruitment and ensuring retention in underserved areas, perhaps through enhanced benefits, better working conditions, or other incentives.

From these findings, we can assume that Indonesia faces a challenge in evenly distributing healthcare professionals, with rural areas experiencing more pronounced shortages. To date, health equity hasn’t been prioritized as a main criterion for evaluating the success of health programs.^51^ Policy interventions to incentivize healthcare professionals to work in underserved areas, scaling up healthcare infrastructure^35^ and education programs to meet the demand in regions with lower healthcare workforce ratios.^32,52,53^

WHO emphasizes the necessity of district-level reporting of health worker data. While provincial-level data offers a general overview, it often fails to represent the situation at the district level. High-level aggregated data can obscure crucial specifics about the distribution and availability of health workers at the subregional level^51^. The strain from a lack of healthcare workers was apparent during the COVID-19 pandemic. Healthcare services almost collapsed during the pandemic peak. Any decrease in healthcare workers led to longer working hours for the remaining staff. This created a domino effect of increasing health workers’ loads^54^. In maternity care, World Bank statistics reveal that maternal mortality rates in Indonesia have persistently remained relatively high from 2017 to 2021 compared to other Southeast Asian countries. A shortage of healthcare professionals is one critical factor influencing the heightened maternal mortality rate in Indonesia^55,56^. NCD demands a workforce for every step, especially preventive care ^57^. Hypertension in Indonesia increased from 25.8% of the population in 2013 to 34.1% in 2018. Notably, many hypertension cases remain asymptomatic, leading to a high prevalence of undiagnosed.

### Limitation

During data collection, we found notable discrepancies between the numbers and distribution of General Physicians and Specialists as reported by the medical association, Medical Council, and Ministry of Health. These inconsistencies likely stem from the council’s reliance on correspondence addresses rather than the actual practice locations of medical doctors. Ministry of Health data is much more focused on public health center workers. We acknowledge limitations in the IDI data, primarily due to a five-year delay from a patient’s death to the data’s release.

## CONCLUSIONS

The imbalance in the distribution of health workers in Indonesia is a burden for this country. Government policy factors, security, economic conditions, individual characteristics, income access to children’s education, knowledge development, and areas with a better socioeconomic scope will be more attractive to health workers, especially specialist doctors.

The health workforce improvement agenda must focus on both increasing the number of health worker and distribute it equally to all people. Central government play a huge role in improving health worker production and distribution to area needed, and Local governments play a vital role in healthcare worker retention. Tailor strategies to local contexts, improve working conditions, offer professional development, and create supportive environments. Engage communities to foster belonging, boosting job satisfaction and retention.

## Data Availability

Data is available by requesting an application through email to the corresponding author.

## ABBREVIATION

HRH: Human Research for Health
HW: Health Worker
NHS: National Health Services
GP: General Practitioner
UHC: Universal Health Coverage

## DECLARATION

### Ethics approval an consent to participate

Not applicable

### Availability of data and materials

Data is available by requesting an application through email to the corresponding author.

### Competiting interests

All authors and contributors to the study stated that they had no conflict of interest.

### Authors’ contributions

**Conceptualization:** Farizal Rizky Muharram, Achmad Chusnu Romdhoni

**Formal analysis:** Farizal Rizky Muharram, Sudhir Anand, Hanif Ardiansyah Sulistya, Fikri Febrian Firmansyah, Julian Benedict Swannjo

**Methodology:** Farizal Rizky Muharram, Hanif Ardiansyah Sulistya, Fikri Febrian Firmansyah, Julian Benedict Swannjo, Muhammad Masrur Rizal

**Project administration:** Alifina Izza, Muhammad Atoillah Isfandiari, Ninuk Dwi Ariningtyas

**Resources:** Farizal Rizky Muharram, Hanif Ardiansyah Sulistya, Fikri Febrian Firmansyah, Julian Benedict Swannjo, Muhammad Masrur Rizal, Alifina Izza

**Supervision:** Achmad Chusnu Romdhoni, Muhammad Atoillah Isfandiari, Ninuk Dwi Ariningtyas

**Writing – original draft:** Farizal Rizky Muharram, Hanif Ardiansyah Sulistya, Julian Benedict Swannjo, Muhammad Masrur Rizal, Alifina Izza

**Writing – review & editing:** Farizal Rizky Muharram, Sudhir Anand, Hanif Ardiansyah Sulistya, Julian Benedict Swannjo, Alifina Izza, Muhammad Atoillah Isfandiari, Ninuk Dwi Ariningtyas

## REFERENCES

1. Jaeger FN, Bechir M, Harouna M, Moto DD, Utzinger J. Challenges and opportunities for healthcare workers in a rural district of Chad. BMC Health Serv Res [Internet]. 2018 Jan 8 [cited 2023 Nov 22];18(1):1–11. Available from: https://bmchealthservres.biomedcentral.com/articles/10.1186/s12913-017-2799-6

2. Jacobs B, Ir P, Bigdeli M, Annear PL, Van Damme W. Addressing access barriers to health services: an analytical framework for selecting appropriate interventions in low-income Asian countries. Health Policy Plan [Internet]. 2012 Jul 1 [cited 2023 Nov 22];27(4):288–300. Available from: 10.1093/heapol/czr038

3. Singh Thakur J, Nangia R, Singh S. Progress and challenges in achieving noncommunicable diseases targets for the sustainable development goals. FASEB Bioadv [Internet]. 2021 Aug 1 [cited 2023 Nov 22];3(8):563–8. Available from: https://onlinelibrary.wiley.com/doi/full/10.1096/fba.2020-00117

4. Leosari Id Y, Uelmen JA, Marc R, Id C. Spatial evaluation of healthcare accessibility across archipelagic communities of Maluku Province, Indonesia. PLOS Global Public Health [Internet]. 2023 Mar 9 [cited 2023 Nov 22];3(3):e0001600. Available from: https://journals.plos.org/globalpublichealth/article?id=10.1371/journal.pgph.0001600

5. Dwi Laksono A, Dwi Wulandari R, Rohmah N, Rukmini R, Tumaji T. Basic Health Survey. BMJ Open [Internet]. 2023 [cited 2023 Nov 22];13:64532. Available from: http://bmjopen.bmj.com/

6. Meliala A, Hort K, Trisnantoro L. Addressing the unequal geographic distribution of specialist doctors in Indonesia: The role of the private sector and effectiveness of current regulations. Soc Sci Med. 2013 Apr 1;82:30–4.

7. Crisp N, Gawanas B, Sharp I. Training the health workforce: scaling up, saving lives. The Lancet [Internet]. 2008 Feb 23 [cited 2023 Nov 22];371(9613):689–91. Available from: http://www.thelancet.com/article/S0140673608603098/fulltext

8. Efendi F, Haryanto J, Indarwati R, Kuswanto H, Ulfiana E, Has EMM, et al. Going Global: Insights of Indonesian Policymakers on International Migration of Nurses. J Multidiscip Healthc [Internet]. 2021 Nov 26 [cited 2023 Nov 22];14:3285–93. Available from: https://www.dovepress.com/going-global-insights-of-indonesian-policymakers-on-international-migr-peer-reviewed-fulltext-article-JMDH

9. Meilianti S, Smith F, Kristianto F, Himawan R, Ernawati DK, Naya R, et al. A national analysis of the pharmacy workforce in Indonesia. Hum Resour Health [Internet]. 2022 Dec 1 [cited 2023 Nov 22];20(1):1–12. Available from: https://human-resources-health.biomedcentral.com/articles/10.1186/s12960-022-00767-4

10. 10. Anderson I, Meliala A, Marzoeki P, Pambudi E. The Production, Distribution, and Performance of Physicians, Nurses, and Midwives in Indonesia: An Update. Health, Nutrition and Population (HNP) Discussion Paper Series [Internet]. 2014 [cited 2023 Nov 22]; Available from: https://ideas.repec.org/p/wbk/hnpdps/91324.html

11. Homepage J, Sutrisno S. Shortages of Medical Doctors in Indonesia, Is It True? Asian Journal of Health Research [Internet]. 2023 Aug 11 [cited 2023 Nov 22];2(2):1–2. Available from: https://a-jhr.com/a-jhr/article/view/121

12. Laksono AD, Ridlo IA, Ernawaty. DISTRIBUTION ANALYSIS OF DOCTORS IN INDONESIA. Indonesian Journal of Health Administration [Internet]. 2020 Mar 28 [cited 2023 Nov 22];8(1):29–39. Available from: https://e-journal.unair.ac.id/JAKI/article/view/14351

13. Indonesia. Law no. 36 of 2014 on Health Care Personnel. 2014. 2014 [cited 2023 Nov 22]; Available from: www.indolaw.org

14. Indonesia. Law no. 17 of 2023 on Health. 2023. [Internet]. 2023 [cited 2023 Nov 22]. Available from: https://peraturan.bpk.go.id/Details/258028/uu-no-17-tahun-2023

15. Direktorat Perencanaan Tenaga Kesehatan KKI. Dokumen Target Rasio Tenaga Kesehatan. 2022;

16. Kumar R, Pal R. India achieves WHO recommended doctor population ratio: A call for paradigm shift in public health discourse! J Family Med Prim Care. 2018;7(5):841.

17. Pandya VS, Sampath N, Yadav R, Mahuli A V, Kapadiya JD. Dental Manpower in India: changing trends upto 2020. Journal of Xidian University. 2021 Jul 3;15(7).

18. The White Ribbon Alliance. A COUNT OF BEDSIDE MIDWIVES IN MALAWI. 2017.

19. World Health Organization. Global strategy on human resources for health: Workforce 2030. 2016.

20. Achim Zeileis, Christian Kleiber. ineq: Measuring Inequality, Concentration, and Poverty [Internet]. 2014 [cited 2022 Dec 2]. Available from: https://CRAN.R-project.org/package=ineq

21. Jin J, Wang J, Ma X, Wang Y, Li R. Equality of Medical Health Resource Allocation in China Based on the Gini Coefficient Method [Internet]. Vol. 44, Iran J Public Health. 2015. Available from: http://ijph.tums.ac.ir

22. Panzera D, Postiglione P. Measuring the Spatial Dimension of Regional Inequality: An Approach Based on the Gini Correlation Measure. Soc Indic Res. 2020 Apr 1;148(2):379–94.

23. Hermawan A. Analisis Distribusi Tenaga Kesehatan (Dokter Perawat Dan Bidan) Di Indonesia Pada 2013 Dengan Menggunakan Gini Index. Buletin Penelitian Sistem Kesehatan. 2019;22(3):167–75.

24. Saito E, Gilmour S, Yoneoka D, Gautam GS, Rahman MM, Shrestha PK, et al. Inequality and inequity in healthcare utilization in urban Nepal: A cross-sectional observational study. Health Policy Plan. 2016 Sep 1;31(7):817–24.

25. Anyangwe SCE, Mtonga C. Inequities in the Global Health Workforce: The Greatest Impediment to Health in Sub-Saharan Africa [Internet]. Vol. 4, Int. J. Environ. Res. Public Health. 2007. Available from: www.ijerph.org

26. Boniol M, McCarthy C, Lawani D, Guillot G, McIsaac M, Diallo K. Inequal distribution of nursing personnel: a subnational analysis of the distribution of nurses across 58 countries. Hum Resour Health. 2022 Dec 1;20(1).

27. Boniol M, McCarthy C, Lawani D, Guillot G, McIsaac M, Diallo K. Inequal distribution of nursing personnel: a subnational analysis of the distribution of nurses across 58 countries. Hum Resour Health. 2022 Dec 1;20(1).

28. Global strategy on human resources for health: Workforce 2030.

29. WHO - Guideline on health policy and system support to optimize community health worker programmes.

30. Global strategy on human resources for health: workforce 2030 Reaffirming the continuing importance of the application of the WHO Global Code of Practice on the International Recruitment of Health Personnel (hereinafter “WHO Global Code”); 2 [Internet]. 2016. Available from: https://sustainabledevelopment.un.org/?menu=1300

31. Bank W, Penh P, Kanchanachitra C, Lindelow M, Johnston T, Hanvoravongchai P, et al. Health in Southeast Asia 5 Human resources for health in southeast Asia: shortages, distributional challenges, and international trade in health services. Lancet [Internet]. 2011;377:769–81. Available from: www.thelancet.com

32. Afenyadu GY, Adegoke AA, Findley S. Improving Human Resources for Health means Retaining Health-Workers: Application of the WHO-Recommendations for the Retention of Health-Workers in Rural Northern-Nigeria. J Health Care Poor Underserved [Internet]. 2017 [cited 2023 Nov 18];28(3):1066–86. Available from: https://pubmed.ncbi.nlm.nih.gov/28804079/

33. Lu H, Hou L, Zhou W, Shen L, Jin S, Wang M, et al. Original research: Trends, composition and distribution of nurse workforce in China: a secondary analysis of national data from 2003 to 2018. BMJ Open [Internet]. 2021 Oct 27 [cited 2023 Nov 18];11(10). Available from: /pmc/articles/PMC8552175/

34. Karan A, Negandhi H, Hussain S, Zapata T, Mairembam D, De Graeve H, et al. Size, composition and distribution of health workforce in India: why, and where to invest? Hum Resour Health [Internet]. 2021 Dec 1 [cited 2023 Nov 18];19(1):39. Available from: /pmc/articles/PMC7983088/

35. Robyn PJ, Shroff Z, Zang OR, Kingue S, Djienouassi S, Kouontchou C, et al. Addressing health workforce distribution concerns: a discrete choice experiment to develop rural retention strategies in Cameroon. Int J Health Policy Manag [Internet]. 2015 [cited 2023 Nov 18];4(3):169. Available from: /pmc/articles/PMC4357984/

36. Hsu YHE, Lin W, Tien JJ, Tzeng LY. Measuring inequality in physician distributions using spatially adjusted Gini coefficients. International Journal for Quality in Health Care. 2016;28(6):657–64.

37. Ismail M. Regional disparities in the distribution of Sudan’s health resources. Eastern Mediterranean Health Journal. 2020 Sep 1;26(9):1105–14.

38. Spatial distribution and associated factors of community based health insurance coverage in Ethiopia: further analysis of Ethiopian demography and health survey, 2019 | BMC Public Health | Full Text [Internet]. [cited 2022 Nov 23]. Available from: https://bmcpublichealth.biomedcentral.com/articles/10.1186/s12889-022-13950-y

39. Zhang T, Xu Y, Ren J, Sun L, Liu C. Inequality in the distribution of health resources and health services in China: Hospitals versus primary care institutions. Int J Equity Health. 2017 Mar 3;16(1).

40. Mirmoeini SM, Shooshtari SSM, Battineni G, Amenta F, Tayebati SK. Policies and challenges on the distribution of specialists and subspecialists in rural areas of Iran. Vol. 55, Medicina (Lithuania). MDPI AG; rrr.

41. Anyangwe SCE, Mtonga C. Inequities in the Global Health Workforce: The Greatest Impediment to Health in Sub-Saharan Africa [Internet]. Vol. 4, Int. J. Environ. Res. Public Health. 2007. Available from: www.ijerph.org

42. Saito E, Gilmour S, Yoneoka D, Gautam GS, Rahman MM, Shrestha PK, et al. Inequality and inequity in healthcare utilization in urban Nepal: A cross-sectional observational study. Health Policy Plan. 2016 Sep 1;31(7):817–24.

43. Laksono AD, Ridlo IA, Ernawaty E. Distribution Analysis of Doctors in Indonesia. Indonesian Journal of Health Administration [Internet]. 2020 Mar 28;8(1):29–39. Available from: https://e-journal.unair.ac.id/JAKI/article/view/14351

44. Heywood PF, Harahap NP. Human Resources for Health Human resources for health at the district level in Indonesia: the smoke and mirrors of decentralization. 2009 [cited 2023 Nov 18]; Available from: http://www.human-resources-health.com/content/7/1/6

45. Samli A, Hadju V, Soma AS. Spatial analysis of health facilities in Mamuju City, West Sulawesi. Enferm Clin [Internet]. 2020 Oct 1 [cited 2023 Nov 18];30:71–5. Available from: https://www.elsevier.es/es-revista-enfermeria-clinica-35-articulo-spatial-analysis-health-facilities-in-S1130862120303570

46. Sumber Daya Manusia K, Sumber Daya Manusia Kesehatan pada Fasilitas Kesehatan Tingkat Pertama dalam Era Jaminan K, Yuyun Yuniar Pusat Penelitian dan Pengembangan Sumber Daya dan Pelayanan Kesehatan dan. Ketersediaan Sumber Daya Manusia Kesehatan pada Fasilitas Kesehatan Tingkat Pertama dalam Era Jaminan Kesehatan Nasional di Delapan Kabupaten-Kota di Indonesia. Media Penelitian dan Pengembangan Kesehatan [Internet]. 2016 [cited 2023 Nov 18];26(4):201–10. Available from: https://www.neliti.com/publications/179261/

47. Rao KD, Shahrawat R, Bhatnagar A. Composition and distribution of the health workforce in India: estimates based on data from the National Sample Survey. WHO South East Asia J Public Health [Internet]. 2016 Sep 1 [cited 2023 Nov 18];5(2):133–40. Available from: https://pubmed.ncbi.nlm.nih.gov/28607241/

48. Meliala A, Hort K, Trisnantoro L. Addressing the unequal geographic distribution of specialist doctors in Indonesia: The role of the private sector and effectiveness of current regulations. Soc Sci Med. 2013 Apr;82:30–4.

49. Anand S. Methods for measuring health workforce inequalities: methods and application to China and India. Human Resources for Health Observer [Internet]. 2010 [cited 2023 Nov 21];5:1–32. Available from: http://www.who.int/hrh/resources/observer/en/

50. Organization WH, Anand S, Fan V. The health workforce in India [Internet]. Geneva: World Health Organization; 2016. (Human Resources for Health Observer Series, 16). Available from: https://iris.who.int/handle/10665/250369

51. Utomo B, Sucahya PK, Utami FR. Priorities and realities: Addressing the rich-poor gaps in health status and service access in Indonesia. Vol. 10, International Journal for Equity in Health. 2011.

52. World Health Organization. Global strategy on human resources for health: Workforce 2030. 2016.

53. Pagaiya N, Kongkam L, Sriratana S. Rural retention of doctors graduating from the rural medical education project to increase rural doctors in Thailand: A cohort study. Hum Resour Health [Internet]. 2015 Mar 1 [cited 2023 Nov 18];13(1):1–8. Available from: https://human-resources-health.biomedcentral.com/articles/10.1186/s12960-015-0001-y

54. Mahendradhata Y, Andayani NLPE, Hasri ET, Arifi MD, Siahaan RGM, Solikha DA, et al. The Capacity of the Indonesian Healthcare System to Respond to COVID-19. Front Public Health. 2021 Jul 7;9.

55. Mahmood MA, Hendarto H, Laksana MAC, Damayanti HE, Suhargono MH, Pranadyan R, et al. Health system and quality of care factors contributing to maternal deaths in East Java, Indonesia. PLoS One. 2021 Feb 1;16(2 February).

56. Sejati EN, Rosa EM, Pramesona BA. Trends and Determinants of the Maternal Mortality Ratio Based on Healthcare Resources. Unnes Journal of Public Health. 2023 Mar 1;12(1):1–11.

57. Suparmi, Kusumawardani N, Nambiar D, Trihono, Hosseinpoor AR. Subnational regional inequality in the public health development index in Indonesia. Glob Health Action. 2018 Dec 3;11(sup1).

